# Viral metagenomic sequencing in a cohort of international travellers returning with febrile illness

**DOI:** 10.1101/2021.05.13.21257019

**Authors:** Alhena Reyes, Ellen C. Carbo, Joost van Harinxma thoe Slooten, Margriet E.M. Kraakman, Igor A. Sidorov, Eric C.J. Claas, Aloys C.M. Kroes, Leo G. Visser, Jutte J.C. de Vries

**Affiliations:** Department of Medical Microbiology, Leiden University Medical Center, Leiden, The Netherlands; Microbiology Department, Hospital Universitario 12 de Octubre, Madrid, Spain; Department of Infectious Diseases, Leiden University Medical Center, Leiden, The Netherlands

**Keywords:** Viral metagenomics, pathogen detection, travellers, fever, capture probes, serum

## Abstract

**Background:** Diagnosis of infections in returning international travellers can be challenging because of the broad spectrum of potential infectious aetiologies potentially involved. Viral metagenomic next-generation sequencing (mNGS) has the potential to detect any virus present in a patient sample and is increasingly being used for difficult to diagnose cases. The aim of this study was to analyse the performance of mNGS for viral pathogen detection in the clinical setting of international travellers returning with febrile illness.

**Methods:** Thirty-eight serum samples from international travellers returning with febrile illness and presenting at the outpatient clinic of the Leiden University Medical Center in the Netherlands in the time period 2015-2016 were selected retrospectively. Samples were processed for viral metagenomic sequencing using a probe panel capturing all known vertebrate viruses. Bioinformatic analysis was performed using Genome Detective software for metagenomic virus detection. Metagenomic virus findings were compared with viral pathogen detection using conventional methods.

**Results:** In 8 out of the 38 patients (21%), a pathogenic virus was detected by mNGS. All viral pathogens detected by conventional assays were also detected by mNGS: dengue virus (n=4 patients), Epstein-Barr virus (n=2), hepatitis B virus (n=1). In addition, mNGS resulted in additional pathogenic findings in 2 patients (5%): dengue virus (n=1), and hepatitis C virus (n=1). Non-pathogenic viruses detected were: GB virus C (n=1) and torque teno viruses (n=3). High genome coverage and depth using capture probes enabled typing of the dengue viruses detected.

**Conclusions:** Viral metagenomics has the potential to assist the detection of viral pathogens and co-infections in one step in international travellers with a febrile syndrome. Furthermore, viral enrichment by probes resulted in high genome coverage and depth which enabled dengue virus typing.

## Introduction

Accurate diagnosis of travel-associated febrile illness in the returning traveller can be challenging, because of the broad spectrum of viral aetiologies potentially involved (1, 2). Identification of potential viral pathogens is important for clinical management and epidemiological reasons.

Metagenomic next-generation sequencing (mNGS) has the potential to detect any known or new pathogen in one single run, in contrast to conventional targeted methods such as PCR. In addition, molecular techniques targeting specific pathogens are dependent upon matching specific primers, leaving variant pathogens unidentified. Finally, emerging pathogens that have not associated with a specific clinical syndrome before, for example encephalitis caused by astroviruses, will be included in the metagenomic width of detection (3).

Since the amount of pathogen in a sample is relatively low and human background is high, several strategies have been applied to increase the sensitivity of mNGS based on physical or enzymatic pre-processing of samples for human DNA depletion (3) (4). Another strategy is hybridization enrichment, with the application of a virome probe panel that targets all known vertebrate viruses to increase the sensitivity of virus detection and characterization (5, 6). Viral enrichment by capture probes increased viral sequence read counts in cerebrospinal fluid samples 100 – 10.000 fold, compared to unenriched sequencing (7). These probe capture panels can also be helpful in the detection of genetic variants and novel viruses up to approximately 40-58% different at nucleotide level from the genome references used in the probe library design (5-8).

Viral mNGS is increasingly being applied directly on different types of samples from patients for pathogen detection in undiagnosed cases, both in retrospective studies (9) and prospective ones (10) with a wide range of proportion of additional findings. A striking example is the rapid and impactful metagenomic analysis of SARS-CoV-2 in 2019 (11). The aim of the current study was to investigate the utility of viral enhanced metagenomic sequencing (mNGS) as a diagnostic tool for viral infections in the returning traveller with febrile illness.

## Material and Methods

### Study design

Retrospectively, a cohort of international travellers with febrile illness upon their return was studied. Patients presenting at the Leiden University Medical Center (LUMC, the Netherlands) from January 2015 to March 2017 with fever after recent international trip and informed consent (12) were enrolled. Serum samples were obtained upon presentation at the first-aid department or outpatient clinic and were tested for dengue antigenemia, malaria, and other infections on clinical suspicion, at the Clinical Microbiology Laboratory of the LUMC as routine diagnostic practice. Patients with proven viral respiratory infections, viral or bacterial gastro-enteritis, or malaria have been excluded from viral metagenomics analysis. Of the included travellers (n=38) serum samples were utilized to perform viral mNGS sequencing independently of conventional test results and diagnosis.

### Ethical approval

Approval was obtained from the ethical committee from the LUMC (P11.165 NL 37682.058.11, and Biobank Infectious Diseases protocol 2020-03 & 2020-04 B20.002).

### Metagenomic next-generation sequencing (mNGS)

The procedure for metagenomic detection using a viral probe capture panel for clinical samples has been validated previously (7). Prior to nucleic acid extraction, serum samples were spiked with fixed amounts of non-human pathogenic viruses as internal sequencing RNA and DNA controls: Equine Arteritis Virus (EAV) and Phocid alpha-herpesvirus (PhHV-1). Nucleic acid extraction was performed using MagNApure 96 DNA and Viral NA Small volume extraction kit on the MagNApure 96 instrument (Roche, Germany), with 200 µl of serum sample input and 100 µl output eluate. STAR Buffer was used as negative control for the entire workflow from nucleotide isolation throughout sequencing. Library preparation was carried out with the NEBNext® Ultra II Directional RNA Library Prep kit for Illumina® and NEBNext® Multiplex Oligos for Illumina® (unique dual index primers pairs, E6440) with 10 ul of pre-concentrated eluate and following a modified version of the protocols for use with “purified mRNA or rRNA Depleted RNA” as described previously (13, 14). Nuclease free water was used as a library preparation control (upstream negative control). Libraries were combined in pools of three libraries from samples plus one negative control for viral capture probe enrichment.

### Viral capture probe enrichment

SeqCap EZ Hypercap probes (Roche), designed to cover the genomes of 207 viral taxa known to infect vertebrates including humans were utilized. A complete list of the viral taxa included can be found in the supplementary tables of the manuscript by Briese et al. (6) The quality and quantity of the amplified libraries before and post-capture were determined using the Agilent 2100 Bioanalyzer (Agilent technologies, Palo Alto, CA, USA) and Qubit (Thermo Fisher, Waltham, MA, USA). For capturing, 250 ng of four amplified DNA libraries were combined in a single pool resulting in a combined mass of 1 ug. For enrichment of the DNA sample library pools, the SeqCap EZ HyperCap Workflow User’s Guide (Roche) was followed with several in-house adaptations to the manufacturers protocol as described previously (7). Subsequently, the target regions were captured by hybridization each pool of four sample libraries with the SeqCap EZ probe pool (6) overnight. The HyperCap Target Enrichment kit and Hyper Cap Bead kit were used for washing and recovery of the captured DNA. Finally, post-capture PCR amplification was performed using KAPA HiFi HotStart ReadyMix (2X) and Illumina NGS primers (5 uM), followed by DNA purification using AMPure XP beads. Final products were sequenced using the Novaseq6000 platform (Illumina, San Diego, California, USA), obtaining up to 10 million of 150bp paired-end reads per sample (GenomeScan B.V. Leiden, the Netherlands).

### Bioinformatic analysis

Primary data analysis, bcl conversion and demultiplexing was performed with bcl2fastq (Illumina). After quality pre-processing, sequencing reads were taxonomically classified and subtyped with metagenomic pipeline Genome Detective (www.genomedetective.com) (15) and classification tool Centrifuge (16) (v1.0.3, GeneBank taxonomy v2019-04-04), including analysis of the proportion of sequence reads assigned to the human genome. The variables collected for virus hits were: number of total, human and viral reads, horizontal coverage (%), number of contigs aligned over the genome, and virus types. Reads were normalized to get the number of reads per Kb genome per Million total reads (RPKM) using the following formula: RPKM= (target reads*1000000*1000)/(total reads after quality check*genome size in base pairs). The following criteria were applied for defining a positive result: horizontal coverage of three or more genome locations without the virus being detected in the negative controls of simultaneous runs. Bacteriophages and human retroviruses known to integrate in human chromosomes were not taken into account. The mNGS detection of a pathogenic virus was subject to confirmatory analysis: RT-PCR on the original sample depending on the virus and reference diagnostic test availability.

### Phylogenetic analysis

Typing and phylogenetic analysis was performed using the Genome Detective Typing Tool (15) for dengue virus.

## Results

### Cohort

Hundred-thirteen returning travellers visited the outpatient clinic of the LUMC from January 2015 to March 2016 with fever. After exclusion of patients with proven respiratory infections and viral or bacterial gastro-enteritis, 38 serum samples from returning travellers with febrile-illness were processed for metagenomic sequencing. The mean age of the 38 travellers was 44,2 years (range 13.3 – 71.9). Seventeen (45%) travellers returned from South or Sub-Saharan African countries, 14 (37%) from Central-Southeast Asia, and seven (18%) from Central-South America.

### mNGS findings in relation to conventional diagnosis

Thirty-eight serum samples were analysed by mNGS. The percentage of sequence reads assigned to the human genome was on average 74% (range 5-96%, data not shown).

Six patients had a viral infection diagnosed by conventional serologic methods during the time of the visit at the outpatient clinic: three dengue virus primary/secondary infections, two Epstein-Barr virus primary infections and one chronic hepatitis B virus infection. All viral infections detected by conventional methods were also been detected by metagenomic sequencing (sensitivity 6/6, 100%). **Table 1** shows the number of reads, genome coverages and dengue genotypes found in relation with the conventional diagnostic test performed. Genome coverage bars are shown in **Figure 1 A**.

**Table 1.** mNGS results from previously diagnosed viral infections. NS1 Ag; non-structural antigen 1, EBV; Epstein-Barr virus, HBV; Hepatitis B virus RPKM; Reads per Kb genome per Million total reads

| mNGS virus finding # | Patient (age range, country or countries visited) | mNGS finding | Conventional positive tests | Diagnosis | # of reads Genome Detective (15) / Centrifuge (16) | # of normalized reads Genome Detective (15) / Centrifuge (16) (RPKM) | Coverage | # of contigs |
| --- | --- | --- | --- | --- | --- | --- | --- | --- |
| 1 | 30-39 y, Brazil | Dengue virus 1<br>Genotype V | NS1 Ag+, IgG+/IgM+ | Dengue virus infection | 824,061/<br>270,449 | 1,172/<br>1,835 | 82% | 11 |
| 2 | 40-49 y, Malaysia | Dengue virus 3<br>Genotype I | NS1 Ag+<br>IgM-/IgG- | Dengue virus infection | 1,050,036/<br>430,486 | 59,994/<br>74,293 | 99% | 1 |
| 3 | 20-29 y, Sri Lanka | Dengue virus 1<br>Genotype I | NS1 Ag+<br>IgM+/IgG- | Dengue virus infection | 173,019/<br>78,729 | 876/<br>989 | 88% | 7 |
| 4 | 40-49 y, South Africa | Human gamma-herpesvirus 4 (EBV) | EBV VCA IgM+/IgG-<br>EBNA IgG -<br>load: 1.020 IU/mL | Primary EBV infection | 463,808/<br>307,178 | 89/<br>69 | 31% | 98 |
| 5 | 20-29 y, Myanmar, Thailand, | Human gamma-herpesvirus 4 (EBV) | EBV VCA IgM+/IgG+<br>EBNA IgG - | Primary EBV infection | 68,999/<br>79,652 | 64/<br>29 | 8% | 43 |
| 6 | 30-39 y, Thailand, Cambodia | Hepatitis B virus | HBsAg+, anti-HBc+,<br>HBe Ag+<br>load: 8.3 log10 IU/ml | Hepatitis B infection, chronic | 206,014,159<br>/61,273,548 | 184,249/<br>311,644 | 100% | 1 |

**Figure 1.**
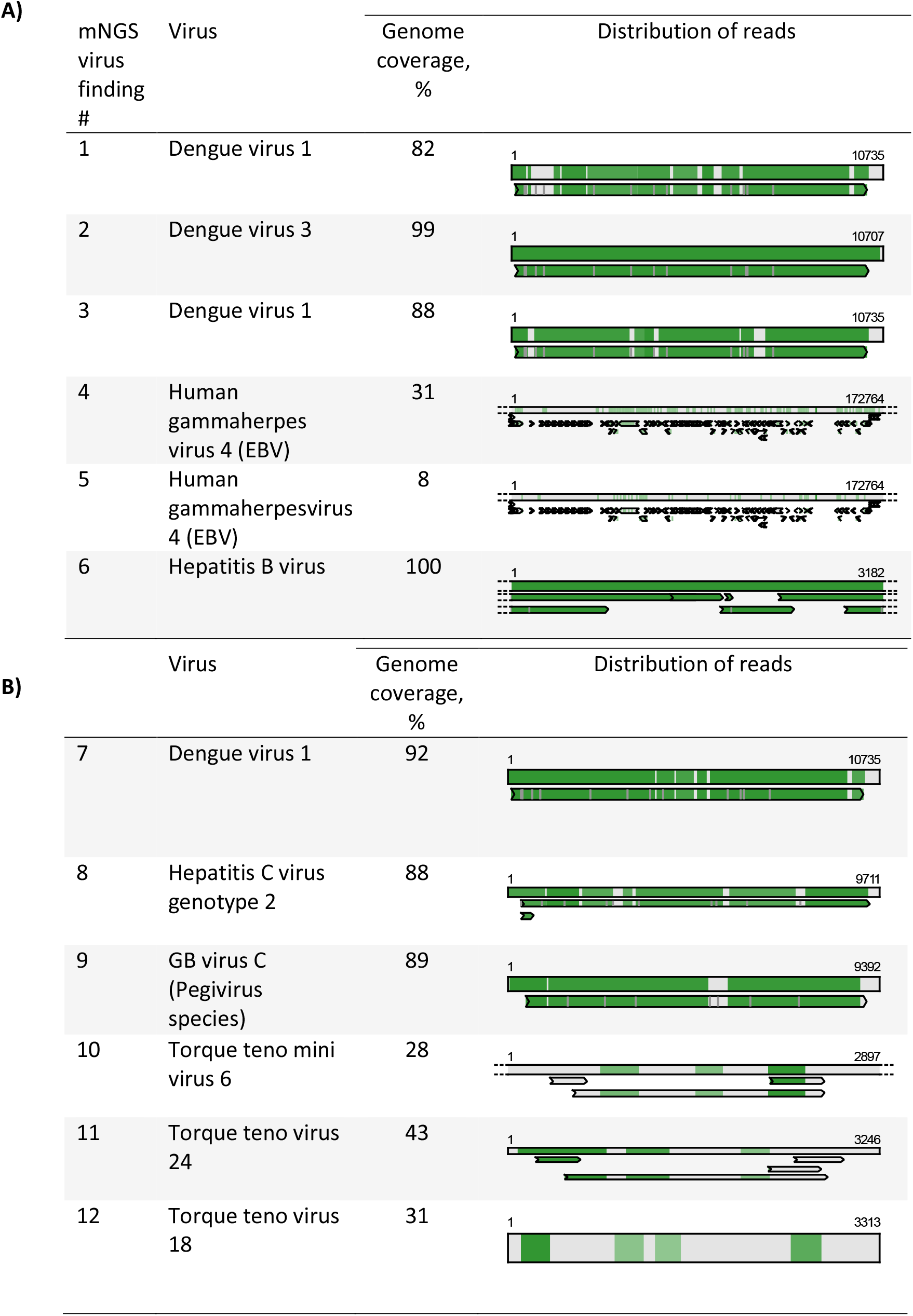
A and B. Horizontal genome coverage of mNGS virus findings in patients with conventional diagnosis (A) and in patients without aetiology by conventional assays (B). Top bar represents nucleotide alignment, bottom bar(s) represents amino acid alignment, green zone: matching sequences. EBV; Epstein-Barr virus

### Additional mNGS findings

Using mNGS, two additional pathogenic viruses were detected (**Table 2, Figure 1 B**): one additional dengue virus and one hepatitis C virus. These infections were confirmed as true positives by qPCR. In the patient in whom the dengue virus infection was diagnosed mNGS, the original dengue NS1 antigen screening test was negative at the time of the visit at the outpatient clinic.

**Table 2.** Additional findings by mNGS, coverage, and confirmatory test results. RPKM; Reads per Kb genome per Million total reads

| mNGS virus finding # | Patient (age range, country or countries visited) | Clinical symptoms | Conventional diagnostics | mNGS finding | # of reads Genome Detective (15) / Centrifuge (16) | # of normalized reads Genome Detective (15) / Centrifuge (16) (RPKM) | Coverage | # of contigs | Confirmatory test results (performed after mNGS) |
| --- | --- | --- | --- | --- | --- | --- | --- | --- | --- |
| 7 | 20-29 y, Indonesia | Fever since day of return, unknown aetiology | Malaria PCR-, smear-, Dengue NS1 Ag-<br>Leptospirosis Ab-/PCR- | Dengue virus 1 genotype IV | 1,718,292/<br>814,613 | 63,303/<br>68,849 | 92% | 6 | Dengue PCR+, Anti-dengue IgG+/IgM-, suggestive for re-infection |
| 8 | 60-69 y<br>Suriname, French Guiana | Fever and joint pain 3d after return, Unknown aetiology | Chikungunya IgG-/IgM-, Dengue IgG+/gM-, Ag-:<br>past dengue infection<br>Blood culture: negative | Hepatitis C virus genotype 2 | 939,937/<br>1,456,226 | 8,820/<br>2,897 | 88% | 7 | Anti-VHC+<br>Load: 5.640 IU/mL<br>NS5B region: Genotype 2 |

The following non-pathogenic viruses were detected: GB virus C (one patient, 1.821.303 sequence reads) and torque teno viruses in three patients: type 6 (147 reads), 24 (892 reads) and 18 (139 reads), coverage bars are represented in **Figure 1B**. The GB virus C was found in co-infection with dengue virus.

### Virus typing

The four dengue virus infections could also be typed and phylogenetic analysis was performed (**Figure 2**). Three of the infections were classified as serotype 1 (genotype I, IV and V) and one as serotype 3 (genotype I). The HCV positive finding was classified as genotype 2 by alignment with reference NC_009823.1 strain.

**Figure 1.**
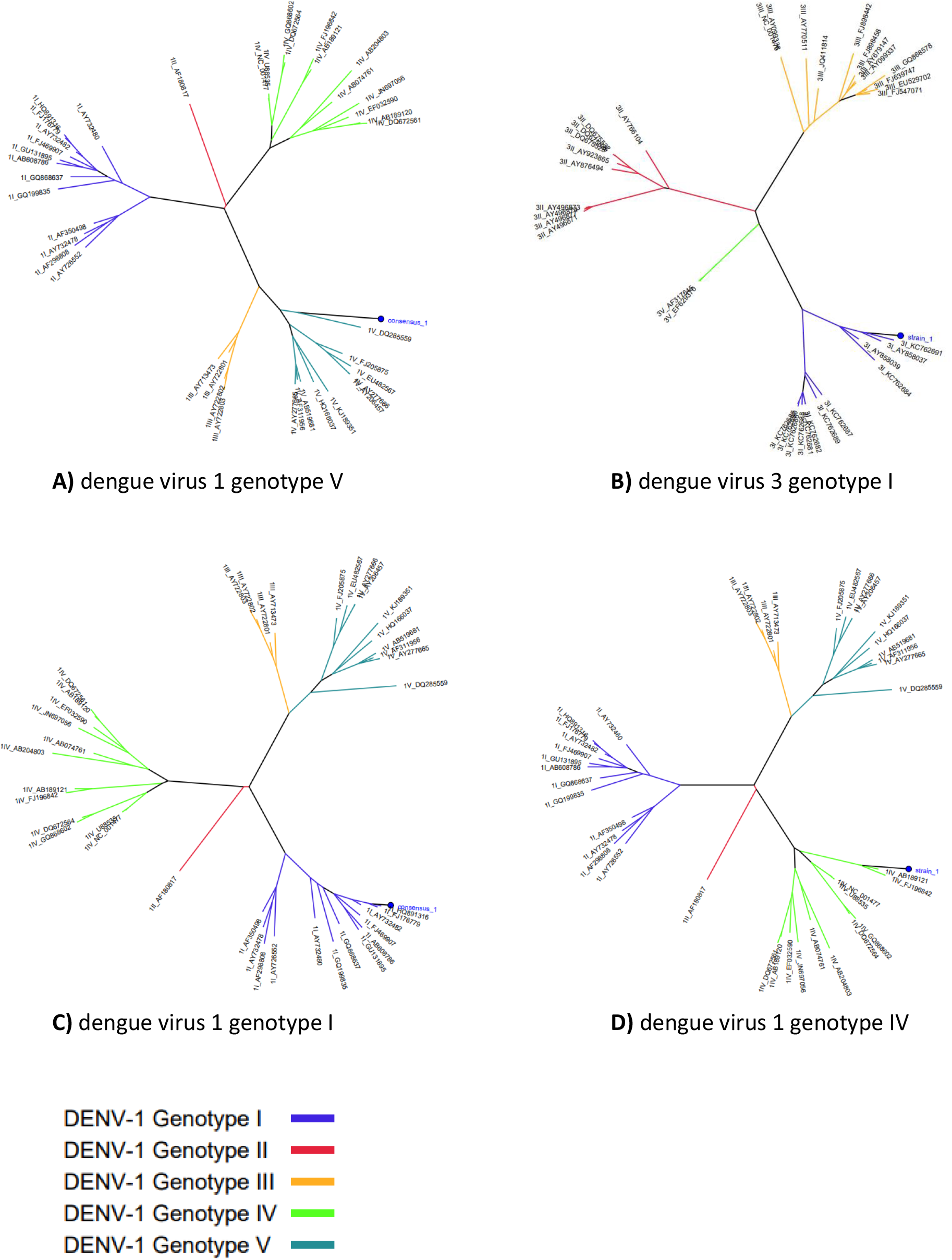
Phylogenetic trees of the dengue viruses detected in the returning travels with fever. The last dengue virus infection was not detected by means of conventional lateral flow assay. The letters correspond to mNGS findings # 1, 2, 3 and 7 respectively in Tables 1 and 2).

## Discussion

In the current study, viral mNGS was successful in detecting six previously diagnosed infections and revealed two new findings (5%) in 38 serum samples from returning travellers with febrile illness. In similar studies using mNGS in returned travellers, new and diverse findings have been reported but none of them used a capture panel for pathogen detection (1, 17). Application of capture probes results in higher coverage of the genomes detected and in more reliable sequences because of an increased sequencing depth (6, 7). As a result, subsequent typing and phylogenetic analysis could be performed using the consensus genome sequences after *de novo* genome assembly.

The diagnosis dengue virus was rejected in one patient after a negative dengue NS1 antigen rapid test upon outpatient visitation, whereas dengue sequences were detected by mNGS and afterwards confirmed by PCR. Dengue virus antigen tests are known for their lower sensitivity after one week of onset of disease and potentially in case of secondary infections with lower loads (18).

Serotypes and subsequent genotypes of dengue virus were available after sequence analysis using a dengue virus typing tool based on *E* gene sequences (1,485bp) for phylogenetical classification. Dengue serotypes differ in more than 30% in their amino acid sequences (19). Dengue virus typing is not relevant for pathogen detection but it is for epidemiological surveillance of infecting strains. Furthermore, dengue virus typing can be of use for differentiating isolates in case of secondary infections, while severity of primary or secondary infections are related to serotypes (20). In our study, all the dengue virus serotypes could be properly identified as a result of increased genome coverage and depth due to effective enrichment.

The detection of hepatitis C virus in serum of a patient highlights the potential of mNGS to diagnose infectious diseases beyond the differential diagnostic list of expected pathogens. The patient was a 60-69 years-old person presenting with fever at the outpatient clinic three days after the return from Suriname and French Guiana. She was tested for dengue antigenemia, and Zika and Chikungunya antibodies which were all negative. There was no suspicion of HCV infection since signs and symptoms were not consistent and transaminases levels were in the range of normality. Although HCV infection is not a cause of febrile illness in the returning traveller, the detection of unsuspected pathogens is crucial for clinical management, therapy administration and prevention of transmission.

GB virus C, formerly known as hepatitis G virus, is a lymphotropic RNA virus of the *Pegivirus* family, it is related to hepatitis C virus and was thought to cause chronic hepatitis in the past, however nowadays is considered non-pathogenic. It has been suggested to have a controversial protective role in HIV-1 infected patients, of which the underlying mechanisms are not fully known (21). A recent study associated GB virus C metagenomic detection with an attenuation of disease caused by Ebola virus through immunomodulation (22). We did not find any report about GBV-C and dengue coinfection or association with dengue pathogenicity. Our finding may rise questions about the possible immunomodulating role of GBV-C virus in dengue when the two viruses are present in blood simultaneously since dengue virus can replicate in lymphocytes.

The use of a virus capture panel has been reported to increase significantly the number of reads and coverages generated in sequencing platforms (7). The high sensitivity of NGS when combined with the viral capture panel does not only enable the finding of clinically unsuspected pathogens, but also may provide a negative predictive value within the range of use for clinical practice (7, 14). The increase in viral sequence reads also enables subsequent typing or detection of potential antiviral resistance. A recent review highlighted the sensitivity of hybridization-based enrichment techniques for viral genomes screening and proposed its deployment as an alternative diagnostic tool when traditional methods fail to detect a pathogen, even when viral genomes differs <40% from probe sequences (23).

In conclusion, the application of viral metagenomics in this study provided the additional detection of two unsuspected viral pathogens and one first report coinfection. Metagenomic sequencing has the potential to diagnose a viral febrile syndrome in the returned traveller with the use of a single test. The use of a broad viral capture panels makes this method more sensitive and generates enough reads and coverages for reliable pathogen identification and typing.

## Data Availability

N/A as data contains human sequence reads, that is not allowed to be shared

